# COVIDNearTerm: A Simple Method to Forecast COVID-19 Hospitalizations

**DOI:** 10.1101/2021.10.08.21264785

**Authors:** Adam B. Olshen, Ariadna Garcia, Kristopher I. Kapphahn, Yingjie Weng, Paul D. Wesson, George W. Rutherford, Mithat Gonen, Manisha Desai

## Abstract

COVID-19 has caused tremendous death and suffering since it first emerged in 2019. In response, models were developed to help predict the course of various disease metrics, and these models have been relied upon to help guide public health policy. Here we present a method called COVIDNearTerm to “forecast” hospitalizations in the short term, two to four weeks from the time of prediction. COVIDNearTerm is based on an autoregressive model and utilizes a parametric bootstrap approach to make predictions. We evaluated COVIDNearTerm on San Francisco Bay Area hospitalizations and compared it to models from the California COVID Assessment Tool (CalCAT). We found that that COVIDNearTerm predictions were more accurate than the CalCAT ensemble predictions for all comparisons and any CalCAT component for a majority of comparisons. For instance, at the county level our 14-day hospitalization median absolute percentage errors ranged from 16% to 36%. For those same comparisons the CalCAT ensemble errors were between 30% and 59%. COVIDNearT-erm is also easier to use than some other methods. It requires only previous hospitalization data and there is an open source R package that implements the algorithm.

## 2 Introduction

Since first being identified in Wuhan, China in 2019, the SARS-CoV-2 virus and accompanying COVID-19 disease has had devastating consequences. As of September 1, 2021, and according to the website worldometers.info, the virus has caused over 4.5 million deaths worldwide and about 650,000 deaths in the United States. At the peak of the pandemic, according to the CDC (https://covid.cdc.gov/covid-data-tracker/#hospitalizations), total hospitalizations have been over 120,000 on a single day in the US. Among those who have recovered many have developed long-term health complications [Blomberg et al., 2021].

To combat the impact of the virus, starting in March and April of 2020 in the United States, various restrictions have been implemented. These restrictions have been maintained to various degrees in different communities. While restrictions were effective at slowing transmission [Wellenius et al., 2021], they have had a large impact on the economy [Pak et al., 2020], which were not separate from the impact of the virus itself [Roy, 2020]. Predictive models, both long-term and short-term, were developed to help inform the level of restrictions and the amount of inpatient medical resources that would be needed.

Among long-term models, the first was from Imperial College [Ferguson et al., 2020]. Here, at the very beginning of the pandemic, long-term predictions were made of healthcare demand that would result from potential interventions. Similar types of long-term predictions have been made throughout much of the pandemic by the Institute for Health Metrics and Evaluation (IHME) [forecasting team, 2020].

In contrast, our model, COVIDNearTerm, is useful for the short term. Its particular use is for “forecasting,” which we define as making predictions two to four weeks into the future. We modeled the near term because we did not believe it was possible to make accurate long term predictions due to the many sources of uncertainty deriving from a once in a century pandemic. We focused on hospitalizations, which typically occur three to ten days after symptoms [Faes et al., 2020], because in the United States they reflect the number of severely ill cases in a population essentially independent of the amount of testing, since almost anyone sick enough is admitted to a hospital. Accurately predicting hospitalizations is particularly important because hospital beds are a limited resource. Using only previous hospitalizations as input, our model’s effectiveness has been validated by comparing predicted hospitalizations to observed hospitalizations.

We are not the first to model short-term hospitalizations. Early in the pandemic Van Wees et al. [2020] developed a Susceptible-Exposed-Infectious-Recovered (SEIR) model for this purpose. Such a model makes predictions through estimating the rate of transitions between these compartments. Perone [2021] compared various time series models to predict near-term hospitalizations in the second wave in Italy. Goic et al. [2021] used an ensemble method to predict short-term ICU hospitalizations.

Our goal was to develop a model that included the major sources of variability in hospitalizations. This included recent trends, typical differences seen between days and random error. To include these factors, we utilized an autoregressive model and made predictions using parametric bootstrap methods. This model can be helpful for answering questions about the future or the past. Regarding the future such a question is what is the expected number of COVID-19 hospitalizations in a county in two weeks? Another example is what is the probability that the number of hospitalizations will exceed a pre-determined threshold, such as 100 in a county, any day in the next two weeks? A question from the past could be did a loosening of restrictions lead to an increase in hospitalizations? For example, four weeks after reopening restaurant dining, did hospitalizations increase more than would be expected given the previous trend?

In June, 2020 the state of California launched the California COVID Assessment Tool (CalCAT, https://calcat.covid19.ca.gov/cacovidmodels/). The purpose was to gather prediction models from the community to help predict the course of the covid pandemic in California in terms of key metrics. An underlying assumption was that by forming an ensemble, a model of models, predictions would be improved. Models include “nowcasts,” the current state of the pandemic, “forecasts”, as we have defined them, and “scenarios,” the long-term impacts of various policies. CalCAT forecasts were used to evaluate the quality of our model.

Our manuscript continues by describing the COVIDNearTerm model for hospitalizations. We then assess how well our model predicts hospitalizations throughout the pandemic in the San Francisco Bay Area. We then compare those results to those deriving from models included in CalCAT. This is followed by Discussion.

## 3 Methods

Our modeling scheme, COVIDNearTerm, relies on an autoregressive model and makes predictions utilizing parametric bootstrap methods.

### 3.1 The basic algorithm

Because COVID-19 cases, and thus hospitalizations, involve exponential growth or decline [Perc et al., 2020], like other infectious diseases [Viboud et al., 2016], we developed a modeling approach able to fit such a pattern. Our basic model to account for this temporal dependency is

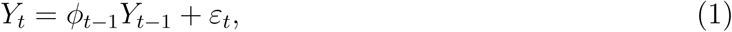

where *Y* represents observed hospitalizations, *ε* represents independent, zero-centered error and *t* represents time in days. The basic idea is that the hospitalizations today are a multiple, represented by *ϕ*, of the hospitalizations yesterday plus random error. For predictions the error is proportional to the value of *Y* as described below.

The training set is the first *T* days of data, and it is on the training data that the parameters of our model are estimated. These parameters are used later to make predictions. We start with a *ϕ* estimate of the form

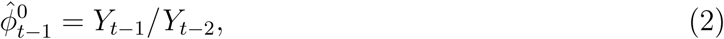

where the 0 superscript represents initial. To smooth daily trends while incorporating local trends the final estimate of *ϕ* is a function of the past *W* days worth of data. The final form is

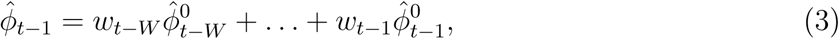

where the *w*s are weights. We considered three weighting schemes. The first was *equal* weights, *w*_*i*_ = 1*/W, i* = 1, …, *W*, and the second was *triangular* weights, 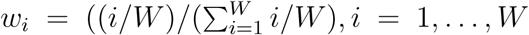. Triangular weights emphasize the most recent observations more heavily, and like equal weights, sum to 1. Note the subscript for *ϕ* of model (1) is *t* − 1, resulting in only data from previous times being used to predict the current time. For our two previous weighting schemes, when a new 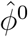 is added, the earliest one within the previous window of length *W* is eliminated. We introduce a third weighting scheme called *unweighted* that is the same as equal weighting except in one way. For unweighted updating we randomly drop one 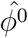 rather than the earliest one, which further damps down recent trends.

Fitting the model to the training set yields distributions 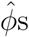 of and 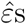. To predict future hospitalizations, we use these distributions and equation (1) to simulate paths of hospitalizations H days into the future, *Ŷ*_*T* +1_, …, *Ŷ*_*T*+*H*_. We go from a measured hospitalization level *Y*_*T*_ to a predicted hospitalization level *Ŷ*_*T*+1_ by simulating 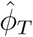 and 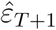, as described below. We repeat this process iteratively to estimate *Ŷ*_*T*+2_, …, *Ŷ*_*T*+*H*_, with each *Ŷ*_*T*+*i*_ based on a previously simulated *Ŷ*_*T*+*i−*1_.

We repeat the simulation process *N* times. From the resulting *N* paths we estimate hospitalizations *H* days into the future by the median of *Ŷ*_*T*+*H*,1_, …, *Ŷ*_*T*+*H,N*_ (or we could use other measures). Alternatively, we can calculate the maximum for each path, *M*_*h*_ = max{*Ŷ*_*T*+1_, …, *Ŷ*_*T*+*H*_}, and estimate the probability of exceeding a trigger within *H* days as the proportion of *M*_*h*_s out of *N* that exceed the trigger.

We derive the estimate 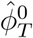 and thus 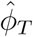 directly from training data including *Y*_*T −*1_ and *Y*_*T*_. For 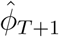 and beyond, we simulate based on our model. We simulate 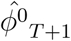 from a 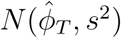 and add that to 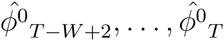 to estimate 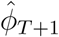 via equation (3). Here 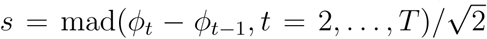, where mad is the scaled median absolute deviation estimate of the standard deviation. We estimate 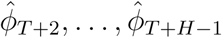 in a similar manner by updating 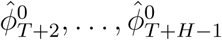.

To simulate 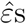, we first model the relationship between *ε* and *Y*. To do this, we use the locally weighted regression method lowess to fit 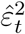 on *Y*_*t*_ for *t* = 1, …, *T*, since this provides an estimate of the variance of *ε* as a function of *Y* based on the training set. We simulate *ε*_*T*+1_, …, *ε*_*T*+*h*_ from a *N* (0, *ν*_*Ŷ*_), where *ν*_*Ŷ*_ is estimated using the value of *Ŷ* (minus the error term) and the lowess fit.

### 3.2 Shrinking the trend estimate

Our experience with COVID-19 hospitalizations time series data is that trends tend to reverse themselves. Increasing trends come down while decreasing trends lead to future increases. Possible causes are that increasing trends lead to more careful behavior or a reversal of opening, while decreasing trends lead to less careful behavior and further opening. We therefore consider shrinking the trend towards *ϕ* = 1. This leads to the shrunken trend model

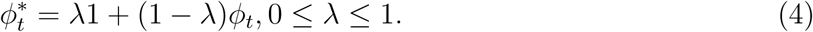

Here *λ* controls the amount of shrinkage, with *λ* = 0 imposing no shrinkage, and thus using the trend exactly as estimated, *λ* = 1 being a model with no trend, and the higher the *λ*, the more the trend is attenuated. The model is utilized by substituting 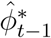 for 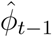 in model (1). The updating of 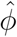 continues as if there was no shrinkage, while 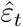 corresponds to the shrunken *Ŷ*_*t*_ (again minus the error term).

To utilize the model we need a method for estimating *λ*. We choose the 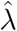 that best fits the training set Ys in the sense of the smallest sum of squared residuals. Alternatively, we could fix 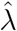 at some value based on previous experience.

### 3.3 CalCAT models

In the Results we include data from nine CalCat models that have been used for hospitalization forecasts at the county level in California. These models are categorized in Table 1, including references to preprints. Further technical details and code for these models can be accessed through CalCAT (https://calcat.covid19.ca.gov/cacovidmodels/). Seven of these models are of the SEIR variety. One model (Simple Growth) is exponential based on the current R-effective and case rate and uses historical hospital and intensive care unit admission rates. Another is a neural forecasting model (UCSB) that assumes predictions can be made by identifying similar patterns across regions from a few months prior. At of the end of the data analysis period on May 1, 2021, only five models remained in CalCAT, four SEIR and Simple Growth. Simple Growth was utilized by CalCAT starting only on December 8, 2020.

**Table 1:**
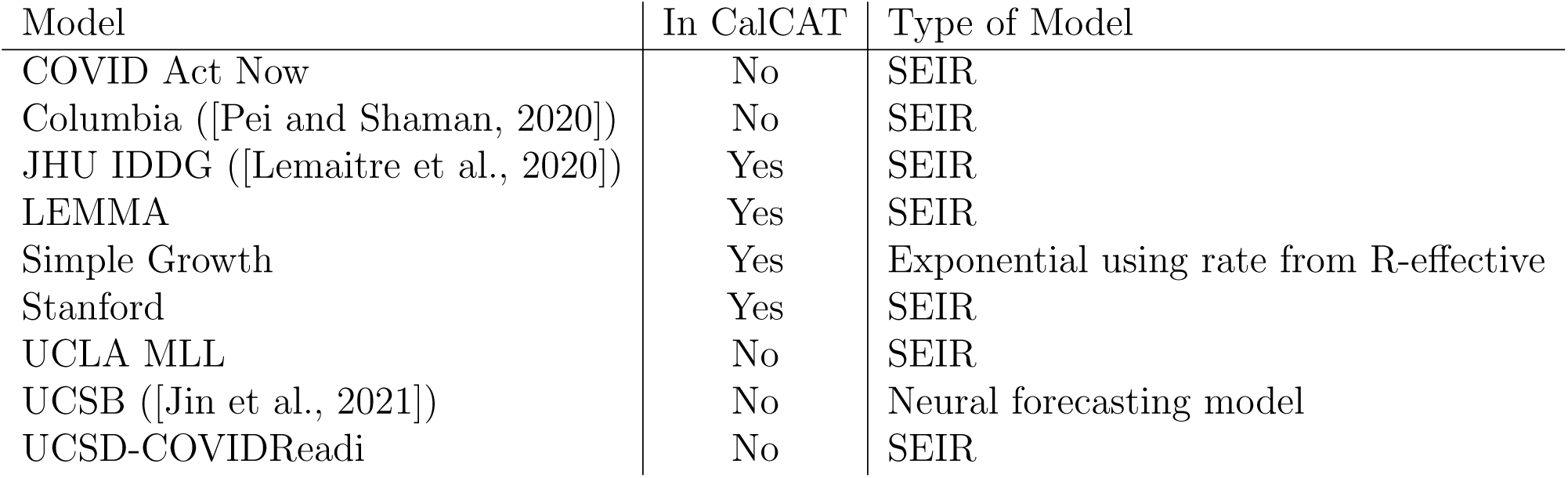
Models used by CalCAT for county-level hospitalization forecasts. Here “No” means the model was once used as part of CalCAT but was not at the end the analysis on May 1, 2021. SEIR stands for Susceptible-Exposed-Infectious-Recovered.

### 3.4 Evaluating Prediction Errors

Because absolute errors in modeling would be expected to increase when hospitalizations increase, we focused on percentage error. More specifically, we utilized the median absolute percentage error (MedAPE). The MedAPE is

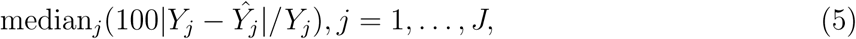

where J is the number of days for which we provided predictions. For some predictions we also report the 25th and 75th percentiles of absolute prediction errors, which we put in brackets. Prediction accuracy was compared between models using the MedAPE.

### 3.5 Data, Code and Utilization

The historical data in this manuscript were downloaded on May 11, 2021 from CalCAT. Code is available at https://github.com/olshena/COVIDNearTerm. It is in the form of an `R` package called **COVIDNearTerm**, and we used version 1.0 for the analysis in this manuscript. Our predictions for all California counties can now be found as part of the CalCAT county forecasts at https://calcat.covid19.ca.gov/cacovidmodels/. While COVIDNearTerm is currently part of the CalCAT ensemble, it was not during the time for which there are comparisons in the manuscript.

The analysis and figures in this manuscript can be reproduced using the file https://github.com/olshena/COVIDNearTerm/Manuscript/reproduce.zip.

## 4 Results

We assessed COVIDNearTerm by comparing it to models in CalCAT. We focused on hospitalization data from the six inner Bay Area counties. They are, ordered by size, Marin (0.3 million people), San Mateo (0.8), San Francisco (0.9), Contra Costa (1.2), Alameda (1.7) and Santa Clara (1.9). We fit our model separately to each county. We started training models with data from May 4, 2020, as we wanted data collection to stabilize before we started, and made our first predictions utilizing data up to June 14, 2020. This gave the first 14-day predictions for June 28, the first 21-day predictions for July 5 and the the first 28 day predictions for July 12. Our final predictions were for May 1, 2021.

We discuss only our models with shrinkage as they had better performance (data not shown). That left us with two parameters to consider: the weighting method and the number of days, *W*, utilized for weighting. As shown in Figure 1, which has the MedAPE for 14-day predictions, the weighting method did not have much impact. For *W*, 14 or 21 tended to do best across counties. Overall, the unweighted method with W=14 gave the smallest MedAPE for Santa Clara (16% [7%,28%]) and San Francisco (23% [10%,45%]), and no other combination was best for more than one other county. Also, across counties, this combination had the lowest average MedAPE of 25.0% [11.4%,42.0%], with the next closest being 25.3% [11.7%,41.8%] for equal weighting with W=14. Therefore, the unweighted method with W=14 was used for all further comparisons.

**Figure 1:**
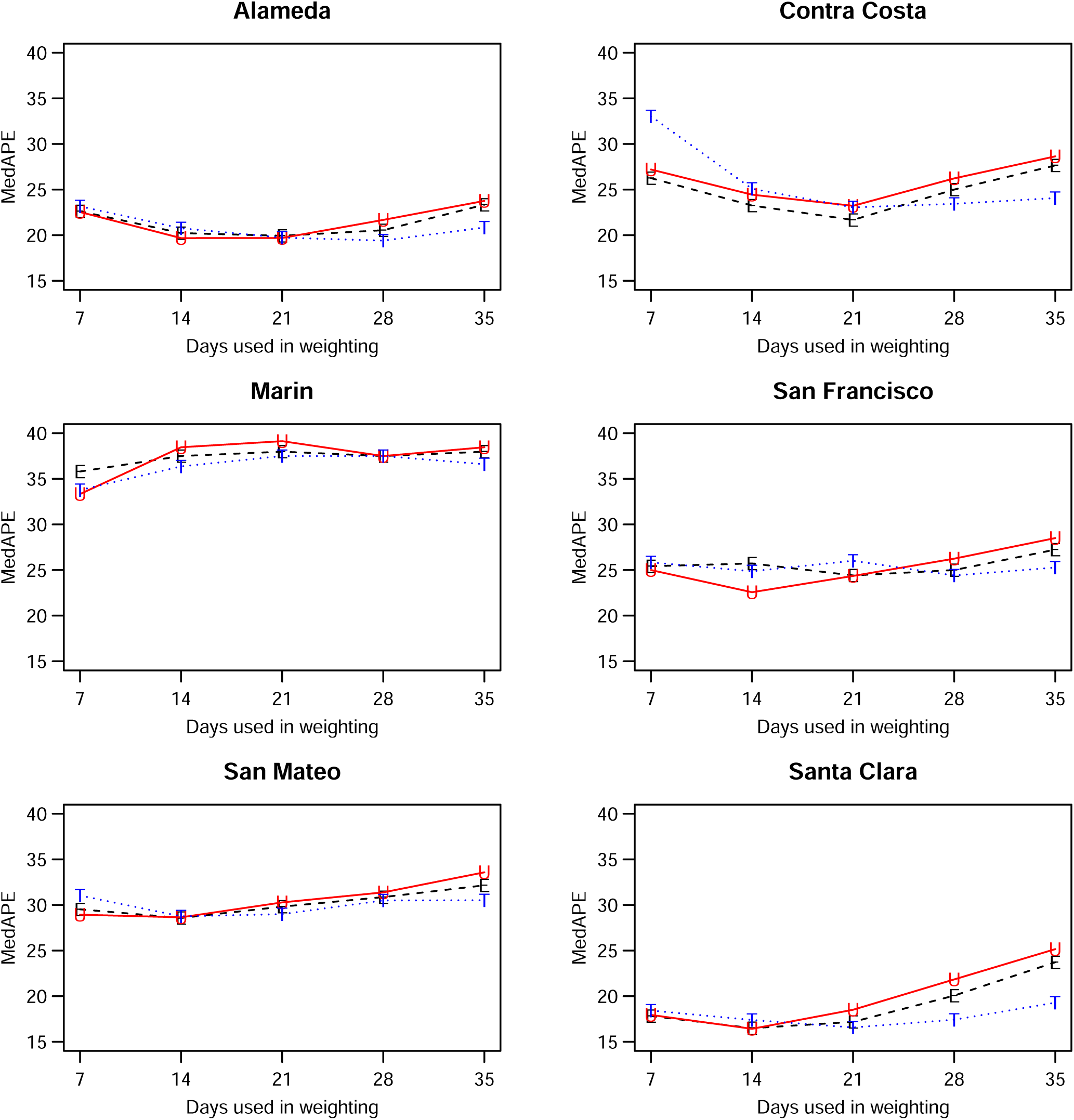
Median absolute percentage error (MedAPE) by county as a function of days used in weighting for 14-day predictions. Here E is for equal (black), U for unweighted (red) and T for triangular (blue).

After selecting our modeling approach, we did further comparisons only on days where CalCAT had ensemble model predictions, which lowered the number of days with predictions from 308 to 283, but changed our prediction errors only slightly. Results across counties for 14-day, 21-day and 28-day predictions can be found in Figure 2 and Table 2. The MedAPEs ranged from 16%-36% for 14-day predictions, 23%-46% for 21-day predictions and 34%-54% for 28-day predictions. All the lowest errors were for Santa Clara (the most populous county) and all the highest errors were for Marin (the least populous county).

**Table 2:** Median absolute percentage error by county for COVIDNearTerm and all CalCAT models at 14 days, 21 days, 28 days. We use the abbreviations AL (Alameda), CC (Contra Costa), MA (Marin), SF (San Francisco), SM (San Mateo) and SC (Santa Clara). The models are discussed in Table 1

| Model | AL | CC | MA | SF | SM | SC |
| --- | --- | --- | --- | --- | --- | --- |
| COVIDNearTerm | 19,28,42 | 25,34,48 | 36,46,54 | 22,38,45 | 27,35,48 | 16,23,34 |
| CalCAT Ensemble | 35,47,65 | 59,81,94 | 51,62,73 | 30,46,58 | 49,61,79 | 31,42,58 |
| COVID Act Now | 68,85,96 | 100,143,170 | 100,100,100 | 99,100,100 | 82,100,100 | 63,81,100 |
| Columbia | 46,54,61 | 83,104,109 | 67,69,85 | 32,46,53 | 63,61,70 | 40,47,52 |
| JHU IDDG | 144,178,202 | 237,301,300 | 320,378,382 | 127,162,180 | 231,301,348 | 53,58,60 |
| LEMMA | 31,41,63 | 28,38,41 | 40,61,66 | 30,43,57 | 35,44,64 | 19,31,42 |
| Simple Growth | 19,28,48 | 88,96,98 | 71,77,83 | 17,19,35 | 66,83,86 | 21,22,29 |
| Stanford | 42,42,45 | 33,34,32 | 60,68,79 | 58,64,78 | 53,56,62 | 50,55,54 |
| UCLA MLL | 38,45,57 | 56,66,76 | 61,67,77 | 50,63,74 | 67,76,81 | 49,59,69 |
| UCSB | 58,77,NA | 57,87,NA | 53,60,NA | 35,43,NA | 54,64,NA | 53,76,NA |
| UCSD-COVIDReadi | 45,55,61 | 70,90,94 | 32,36,46 | 35,36,40 | 35,44,56 | 39,51,67 |

**Figure 2:**
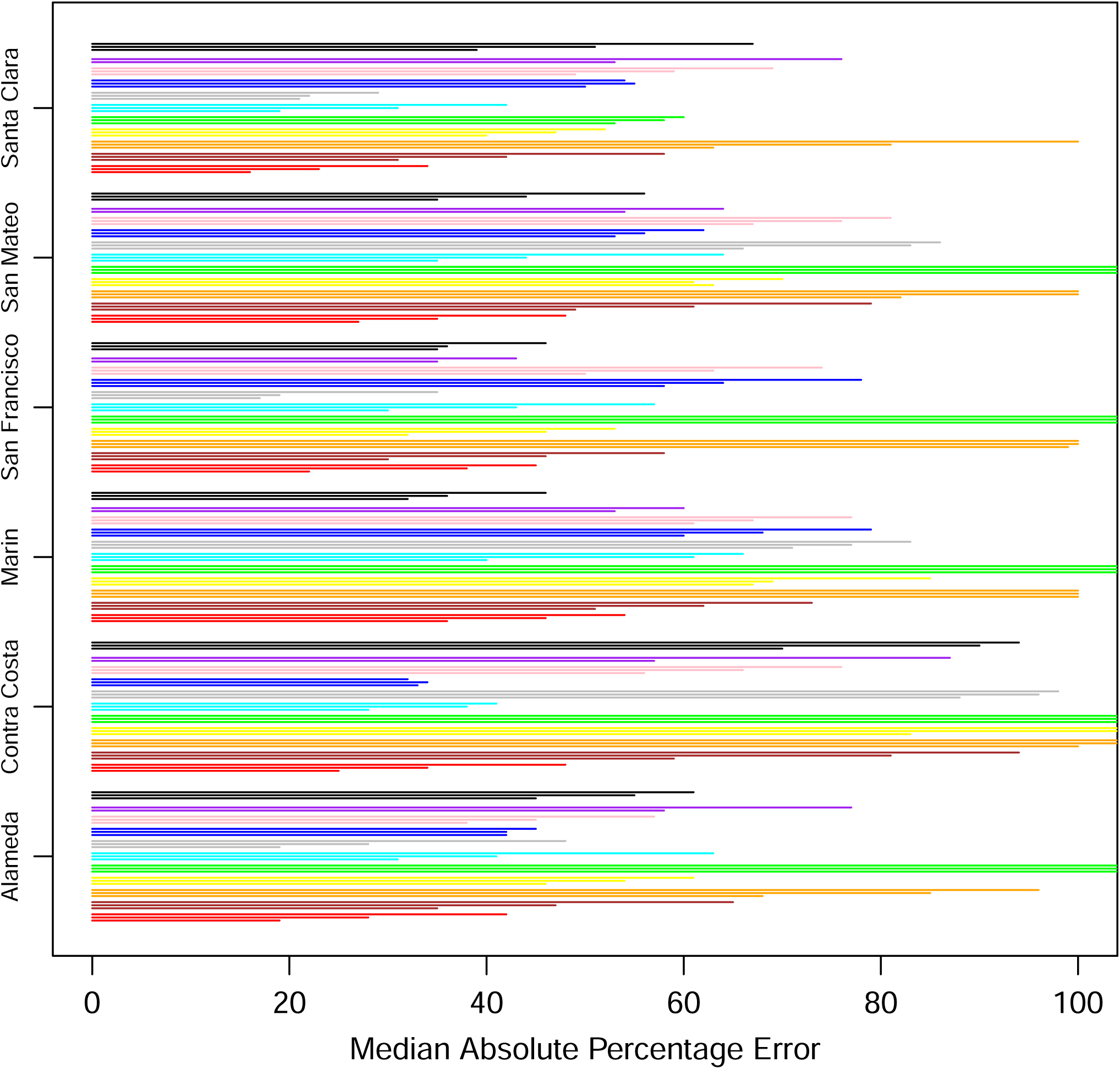
Median absolute percentage error by county for 14 days (bottom of trio), 21 days (middle of trio) and 28 days (top of trio). The models from bottom to top are COVIDNearterm (red), CalCAT Ensemble (brown), COVID Act Now (orange), Columbia (yellow), JHU IDDG (green), LEMMA (cyan), Simple Growth (gray), Stanford (blue), UCLA MLL (pink), UCSB (purple), and UCSD-COVIDReadi (black).

We compared our results to those from the CalCAT tool. The comparisons based on the MedAPE were to both the CalCAT Ensemble and the individual CalCAT components. COVIDNearTerm was more accurate than the Ensemble for all counties at 14, 21 and 28 days. For instance, in Santa Clara County, the COVIDNearTerm MedAPEs were 16% [7%,26%], 23% [10%,47%] and 34% [15%,67%] at 14, 21 and 28 days compared to 31% [14%,49%], 42% [24%,65%] and 58% [31%,84%] for the Ensemble. For Alameda, the MedAPEs for the same comparisons were 19% [11%,30%], 28% [13%,45%] and 42% [16%,65%] versus 35% [17%,59%], 47% [26%,78%] and 65% [46%,102%].

The 14-day predictions for COVIDNearTerm, the CalCAT Ensemble, and two promising methods, LEMMA and Simple Growth, can be seen in Figure 3. Qualitatively, COVIDNearTerm had better predictive performance than the Ensemble in part because it more quickly predicted a decrease after the January 2021 peak. Differences among COVIDNearTerm and LEMMA and Simple Growth were less systematic.

**Figure 3:**
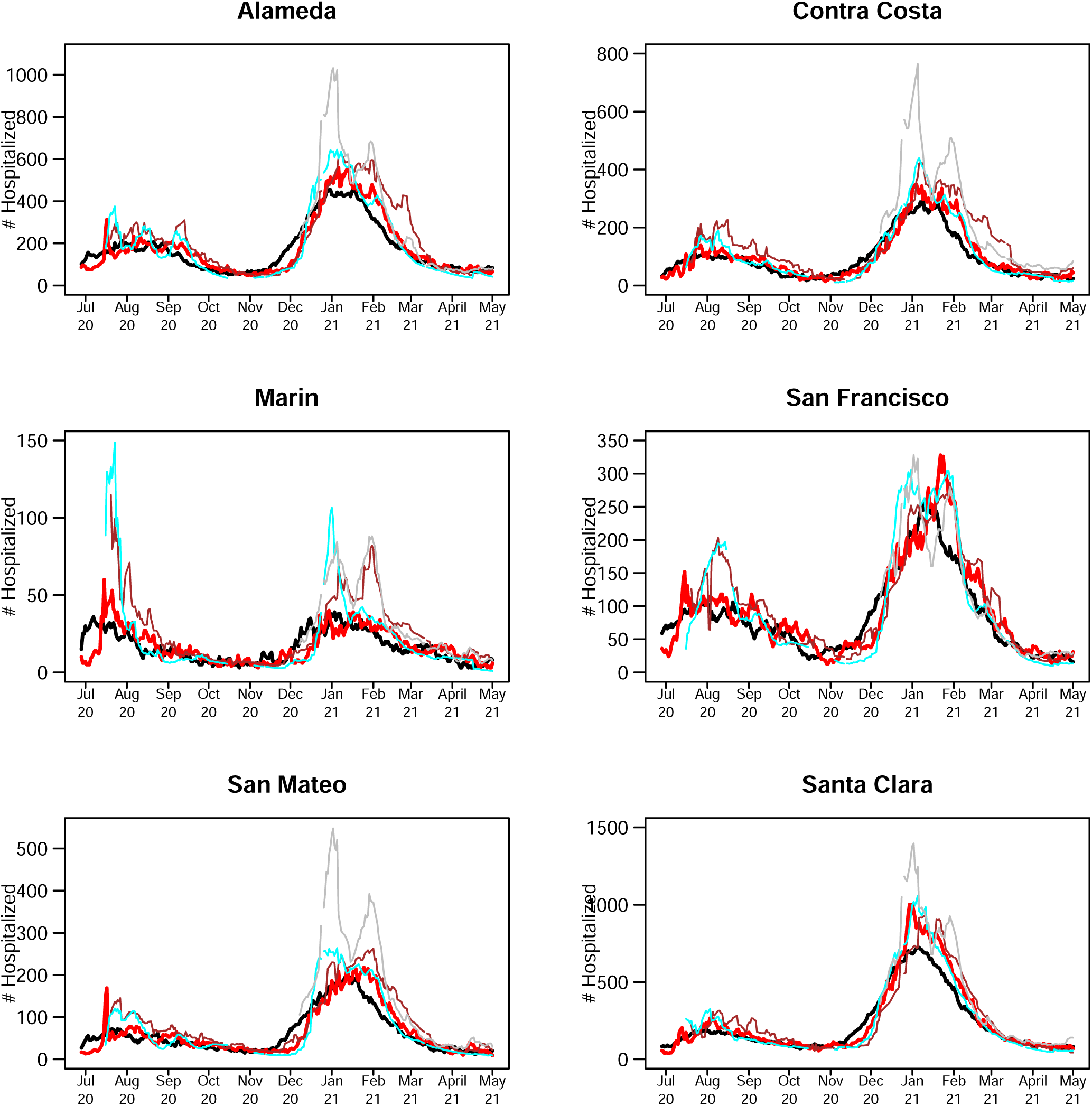
14-day predictions for multiple methods. The lines are for truth (black), COVIDNearTerm (red), CalCAT Ensemble (brown), LEMMA (cyan) and Simple Growth (gray). Note that Simple Growth was utilized by CalCAT starting only on December 8, 2020.

At the individual model level, COVIDNearTerm was most accurate except where stated. For Alameda, Simple Growth was equally accurate for 14-day predictions (MedAPE=19%) and 21-day predictions (MedAPE=28%). For Contra Costa, Stanford was equally accurate for 21-day predictions (MedAPE=34%) and more accurate for 28-day predictions, with MedAPE=32% versus 48% for COVIDNearTerm. LEMMA was also more accurate for 28-day predictions with MedAPE=41%. UCSD-COVIDReadi was more accurate for Marin at 14 21 and 28 days, with MedAPEs of 32%, 36% and 46% versus 36%, 46% and 54% for COVIDNearTerm. For San Francisco, Simple Growth was more accurate for all three with MedAPEs of 17%, 19% and 35% versus 22%, 38% and 45% for COVIDNearTerm. UCSD-COVIDReadi was also more accurate for 21-day (MedAPE=36%) and 28-day (MedAPE=40%) predictions. For Santa Clara, Simple Growth was more accurate for 21-day and 28-day predictions, with MedAPEs of 22% and 29% versus 23% and 34% for COVIDNearTerm.

Overall, COVIDNearTerm was most accurate or tied for most accurate for 10 of 18 comparisons and four of six 14-day comparisons, all but Marin and San Francisco. It was also never worse than the third most accurate model for any comparison.

## 5 Discussion

We developed an autoregressive model called COVIDNearTerm for forecasting hospitalizations two to four weeks out. It has the virtue of only requiring previous hospitalization data, so it is widely applicable. When applied to CalCAT data, its predictions were more accurate than the CalCAT ensemble model for all comparisons and more accurate than any other CalCAT model for most comparisons.

As mentioned, COVIDNearTerm uses only past hospitalizations to predict future hospitalizations. This approach would be disadvantageous if the predictions were inferior to those based on other factors such as testing, community transmission and the percentage of people who have already been infected. We have demonstrated, however, that COVIDNearTerm is as accurate or more accurate than models that include such covariates. Therefore, we were able to do more with less.

There are a few caveats to our modeling. Our comparisons to the models in COVIDNearTerm may have been slightly biased to favor COVIDNearTerm. First, we know the publication date of the CalCAT component models, which is when the predictions appeared on the CalCAT site, but not the actual date the predictions were made. We assume that the publication date was close to the prediction date, and the modelers did have the option of making frequent prediction updates. Second, we looked at the data retrospectively with COVIDNearTerm. Therefore, we had the most updated data on actual hospitalizations, which might be slightly different than when the other models made their predictions. But since we did not start our comparisons until June 2020, most data issues should have been resolved. Overall, we believe the performance metrics we have presented are valid.

We saw a modest impact on hospitalizations based on the day of the week, which was much less than the impact of day of the week on cases (data not shown). If adjusting for day of the week is desired, we suggest making the adjustment outside of COVIDNearTerm. By making predictions 14, 21 or 28 days in the future we mostly avoided the day of the week problem. We also did not address the modest impact of holidays.

One weakness of COVIDNearTerm is that its predictions generally monotonically increase or decrease over time. We might believe that hospitalizations will, for instance, decrease four weeks out based on a recent decrease in cases or test positivity, even if this reduction has not yet been seen in hospitalizations. For this situation, the next generation of COVIDNearTerm could include an adjustment for covariates. How to adjust, however, is not straightforward, as the relationships among covariates and hospitalizations can change over time (data not shown).

There are a couple additional details to discuss when using COVIDNearTerm. First, our model is appropriate for outcomes that exponentially increase or decrease and that start one time period (such as day) where they ended the previous time period. It would be less appropriate for outcomes where measurements are independently measured in each time period, such as new hospitalizations. Second, users of COVIDNearTerm may want to optimize over weighting schemes and numbers of days that go into weighting, that is *W*.

We have shown that a simple hospitalization modeling strategy was as effective as anything else used by CalCAT at a time of great uncertainty. We believe that the less you know at the time of modeling the more a simple approach makes sense. Further, our modeling strategy can be extended to other contexts. For example, it may be helpful in some places in the world where related covariates are missing or not measured accurately. It might also be applicable to other pandemics, including ones resulting from new SARS-CoV-2 variants.

## Data Availability

All data produced are available online at https://github.com/olshena/COVIDNearTerm/Manuscript/reproduce.zip

## Acknowledgments

This work was supported in part by a Rise Grant from Stanford ChEM-H (to M.D.) and NCI P30CA008748 (to M.G.).

